# Investigating dietary quality among individuals aged 15 years and over by diabetes status in South Africa

**DOI:** 10.1101/2024.10.25.24316103

**Authors:** Matthew Burgess, Nuala McGrath

**Affiliations:** CHERISH programme, School of Primary Care, Population Sciences and Medical Education, Faculty of Medicine, University of Southampton, Southampton, UK; Department of Social Statistics & Demography, Faculty of Social Sciences, University of Southampton, Southampton, UK; Africa Health Research Institute, Durban, KwaZulu-Natal, South Africa

**Keywords:** Type 2 Diabetes, Diet, South Africa, Fruit and Vegetables, Sub-Saharan Africa, Ordered Logistic Regression

## Abstract

Analysis of the 2016 South African Demographic and Health Survey (SADHS) estimated that 11.7% of individuals aged 15+ years had poor glycaemic control, despite only 4.7% reporting a previous diabetes diagnosis, and a further 64.5% had prediabetes. Diet-related lifestyle change has an important role in diabetes prevention and management, however entrenched racial and socioeconomic inequalities and increasing urbanisation may present barriers to a healthy diet.

Using data from the 2016 SADHS we investigated whether dietary choices differ by diabetes status defined by previous diagnosis and survey HbA1c, and whether the diet of people living with diabetes (PLWD) differs by age, gender, ethnicity and wealth quintile. Reporting of fruit, vegetable, sugar-sweetened beverage, fruit juice and fast-food consumption was used to create an index of healthy diet. Ordered logistic regression modelling considering the proportional odds assumption was used to investigate the effect of diabetes status and sociodemographic status on healthy diet among the general population and PLWD.

Concurrent low consumption of fruit, vegetables and fruit juice was the most common dietary pattern among both the general population and people living with diabetes, with high consumption of fast-food and sugar-sweetened beverage less common. Among the general population, previous diabetes diagnosis, age ≥55 years, non-black African ethnicity and being in the wealthiest quintile were significantly associated with increased odds of a healthier diet. Among PLWD, there was no association between previous diabetes diagnosis and healthy diet, high wealth remained significantly associated with a healthier diet, whilst female gender and having health insurance also became significantly associated with a healthier diet.

Future public health interventions should focus on making fruit and vegetables more accessible to younger, black and socioeconomically poor populations, irrespective of diabetes status.

## Introduction

An epidemiological transition is currently ongoing in many low- and middle-income countries (LMICs), with infectious diseases such as HIV, malaria and tuberculosis decreasing as leading causes of mortality and being replaced by non-communicable diseases (NCDs), such as type two diabetes mellitus (T2DM)^1^. T2DM prevalence in South Africa has increased during the previous decade, from 7.1% in 2011 to 10.8% in 2021^2^, with as many as 60% of cases undiagnosed^3^. This undiagnosed population is especially vulnerable to the micro and macrovascular complications of T2DM including heart attack, stroke, renal failure and retinopathy. Also vulnerable are those with a diagnosis of T2DM who have not received treatment or whose diabetes remains uncontrolled despite receiving treatment, although this is a considerably smaller group compared to the undiagnosed population^3^.

T2DM incidence and progression is strongly associated with obesity^4^ and interventions in high-income countries such as individualised dietary advice^5^ and diets low in fat and refined carbohydrates^6^ have been shown to improve glycaemic control and prevent T2DM complications. These however are resource intensive and do not consider cultural differences around diet, so are less generalisable to LMICs, and there is relatively little evidence trialling both individualised and population-level dietary interventions to reduce T2DM incidence and progression in South African adults. The prevalence of overweight and obesity in sub-Saharan Africa has risen over the past three decades and South Africa has the highest obesity rates in the region^7^. These changes, coupled with poor diabetes screening and surveillance in South Africa, leaves many people vulnerable to T2DM and its complications.

Despite its classification as an upper-middle-income country^8^, the legacy of colonialism and the devastating apartheid regime means that South Africa has some of the highest rates of socioeconomic and racial inequality globally^9^, which are also reflected in healthcare^3^. Black South Africans have been shown to have poorer dietary diversity than white South Africans and consume energy-dense foods from informal vendors more frequently^10^. These findings fall among a wider backdrop of poor dietary diversity, with the nationally representative SANHANES-1 study reporting a low intake of fruit and vegetables (two or fewer portions of either) in a quarter of South Africans^11^. Internal migration may also have an impact on diet. South Africa now has the most urbanised population in sub-Saharan Africa, with rural-urban migration resulting in 62% living in cities^12^. Globally, both urban residence and lifetime exposure to urban environments have been associated with higher incidence of T2DM and being overweight^13^, whilst studies in urban and peri-urban South Africa have documented dietary composition changing towards the ‘western diet’^12^ - a high intake of energy-dense and nutrient-poor foods and drinks with a low intake of nutrient-dense fresh fruit, vegetables, lean meat and fish. The obesogenic environment of many urban areas may therefore predispose to the dual risk of obesity and food insecurity, with intake of energy-dense foods and features of food insecurity such as childhood stunting seen concurrently within neighbourhoods and households^14^, creating an additional challenge for healthcare providers and policymakers.

The 2016 South African Demographic and Health Survey (SADHS)^15^ is a large nationally-representative survey that additionally collected biomarker samples alongside survey data on a range of health and social outcomes. The 2016 SADHS estimated that 13% of women and 8% of men aged over 15 years had poor glycaemic control (glycosylated haemoglobin (HbA1c) ≥ 6.5%), despite only 5% of women and 4% of men reporting a previous diagnosis of diabetes at the time of survey. Biomarker sampling also showed that 64% of women and 66% of men aged over 15 years had prediabetes (5.7 ≤ adjusted HbA1c ≤ 6.4)^15^, indicating that a large proportion of the population are at risk of developing T2DM. Despite this, few statistical analyses of the associations between diabetes status and diet in South Africa have been conducted, and most studies conducted are highly localised or population specific. More extensive, biomarker-focused, analysis of population surveys such as the SADHS may shed more light on dietary differences between sociodemographic groups in relation to T2DM and inform the design of future public health interventions. This study aims to investigate whether the odds of a healthy diet differs by diabetes status when controlling for important sociodemographic variables (research question one), as well as investigating the association between sociodemographic factors and healthy diet among people living with diabetes (PLWD) (research question two).

## Methods

### Data Collection

Cross-sectional survey and biomarker data from the 2016 SADHS was used with this data collected between 27th June and 4th November 2016. The survey was administered by Statistics South Africa and the South African Medical Research Council. The sampling frame for the SADHS was created using 2011 census enumeration areas (EAs), divided into Primary Sampling Units (PSUs)^15^. The sampling frame contains information about the geographic type and the estimated number of residential dwelling units (DUs) in each PSU. The SADHS 2016 followed a stratified two-stage sample design with probability-proportional-to-size sampling of PSUs at the first stage and systematic sampling of DUs at the second stage^15^. Women aged over 15 years in odd-numbered DUs were eligible for the individual questionnaire, whilst both men and women aged over 15 years in even-numbered DUs were eligible for the individual questionnaire. Participants gave informed verbal consent for interview which was witnessed by the interviewer and documented in the questionnaire. Participants aged over 15 years (with no upper age limit) in even-numbered DUs were eligible for biomarker measurement, including HbA1c, with written consent. For participants aged 15-17, consent was required from both the participant and their legal parent/guardian. HbA1c was measured using dried blood spot sampling with HbA1c ≥ 6.5% indicating diabetes, 5.7 ≤HbA1c ≤ 6.4 prediabetes and HbA1c ≤ 5.6 no diabetes, consistent with WHO^16^ and DHS classifications^15^. These HbA1c classifications were used to define diabetes status in all analyses, are adjusted for sample type and referred to as ‘HbA1c’ for the remainder of this paper. Further information can be found in the SADHS 2016 report^15^. Data was first accessed by the authors on the 20^th^ December 2021 and the authors did not have access to any information that could identify individual participants at any stage of the analysis.

Participant-reported healthy diet group was our categorical outcome measure, with this composite outcome reflecting extent of healthy diet and calculated from reports of five dietary choices: consumption of fast-food, fruit, vegetable, sugar-sweetened beverage (SSB) and fruit juice. The associated 2016 SADHS survey questions, survey answer options and recoding of options into categories for analyses are contained within table 1. The coding of fruit and vegetable consumption as high = >2 types/day reflects the national recommendation of five daily portions of fruit and vegetables^17^ (with fruit juice as the 5^th^ portion).

**Table 1.** Coding of variables comprising the healthy diet group variable and their corresponding SADHS survey question.

| Variable | Survey Question | Answer Options | Considered in Analysis as |
| --- | --- | --- | --- |
| Fast food consumption | How often do you eat fast-foods or take-away foods from places like Chicken Licken, KFC, Captain DoRego's, Steers, Nando's, McDonalds, pizza delivery, etc? | 'Every day', 'at least once a week', 'occasionally', 'never'. | Binary (high if every day or at least once a week, low if occasionally or never) |
| Fruit consumption | Yesterday, how many types of fruit did you eat? | Continuous variable | Binary (high if $\geq 2$, low if $< 2$) |
| Vegetable consumption | Yesterday, how many types of vegetables, excluding potatoes, did you eat? | Continuous variable | Binary (high if $\geq 2$, low if $< 2$) |
| Sugar-sweetened beverage consumption | Yesterday, did you drink any sugar-sweetened drinks?<br>Sugar-sweetened drinks include fizzy drinks like Coke or drinks like Squash where water is added, but not diet or unsweetened cold drinks. | Yes/No | Binary |
| Fruit juice consumption | Yesterday, did you drink any fruit juice? | Yes/No | Binary |

Healthy choices are high fruit, high vegetable, high fruit juice, low fast-food and low SSB consumption. Globally, evidence shows that increased fruit and vegetable consumption reduces risk of cardiovascular diseases^18^ that are also associated with T2DM, whilst studies of more resource-intensive interventions have shown that prescribing a diet high in fruit and vegetables and low in fat and refined sugar resulted in a significant reduction in fasting glucose and HbA1c^19^. Unhealthy choices are low fruit, low vegetable, low fruit juice, high fast-food and high SSB consumption. SSBs have a significant dose-dependent association with obesity and T2DM^20^ whilst fast-food consumption is positively associated with weight gain and obesity^21^. Research into the impact of ultra-processed foods (UPFs) on health also showed that SSBs and industrially prepared meals are among the leading sources of UPFs, which are collectively associated with an increased incidence of T2DM and increased obesity prevalence^22^. Despite being included in the SADHS, interest in lowering salt consumption was excluded from this study due to not being a direct indicator of salt consumption, whilst fried food consumption was excluded due to its similarity to fast-food consumption.

Individuals were initially categorised into one of six groups based on their number of healthy dietary choices (ranging from five healthy, zero unhealthy to zero healthy, five unhealthy). As there were few individuals with zero or five healthy choices, the zero healthy choice and one healthy choice groups were combined to create a single unhealthy diet group and the four healthy choices and five healthy choices groups were combined to form a single healthy diet group. The two healthy choices and three healthy choices groups were retained independently as ‘somewhat healthy’ and ‘moderately healthy’ groups respectively, creating a four-group variable of the extent of healthy diet.

Body mass index (BMI), doctor’s diagnosis of heart attack, doctor’s diagnosis of stroke and doctor’s diagnosis of diabetes were chosen as potential individual confounding variables, on the rationale that recognition of high BMI and NCD-related diagnoses are opportunities to initiate lifestyle change^23^. Additionally, as being diagnosed with diabetes itself is an opportunity to initiate dietary change and good glycaemic control, a joint variable combining diabetes status by HbA1c and previous diagnosis was created to differentiate individuals with ‘controlled diabetes’, i.e. a diagnosis of diabetes, but an HbA1c<6.5% at time of survey. Participants who reported that they did not know if they had ever been diagnosed with diabetes (N=31) were excluded from analyses. Participants who had not had height and weight measured for BMI to be calculated were retained in analyses provided they had a valid HbA1c, with these individuals represented in models as a separate ‘not recorded’ BMI category. Age group^24^, gender^24^, ethnicity^11^ and type of place of residence (urban/rural)^11,25^ were included as known demographic risk factors for healthy diet grouping and T2DM status. Highest level of education completed^26^, wealth quintile^26^, employment in past 12 months^26^ and health insurance coverage^27^ were included as potential socioeconomic confounding variables. Demographic and socioeconomic variables were chosen based on their association with healthy diet or diabetes status in existing literature. Given the limited number of participants reporting doctor’s diagnosis of heart attack and doctor’s diagnosis of stroke, these reports were combined to create a single doctor’s diagnosis of heart attack and/or stroke variable, whilst age was recoded into three age categories: younger adults (15-34 years), middle-aged adults (35-54 years) and older adults (55 years and over).

### Statistical Analysis

All analyses were carried out in Stata Standard Edition 17.0 for Windows (StataCorp, 2022). Descriptive analyses used two-stage sampling weights that accounted for the sampling design. To visualise clustering of dietary choices within healthy diet groups, we used the UpSetR (v1.4.0) package^28^ in R. Patterns were explored overall and for both genders independently. Two-way tables with Chi-squared tests were used to investigate the initial association between potential confounding variables and healthy diet group.

As our healthy diet outcome had four ordered categories, we initially used ordered logistic regression models for the first research question to explore how diabetes status defined by our joint diabetes status variable was associated with healthy diet. Bivariate analysis explored the ‘unadjusted’ association of the joint diabetes variable and each other variable of interest with healthy diet. However, Brant testing identified that several (but not all) of the considered variables included in bivariate analysis violated the proportional odds assumption (shown in appendix 1). Thus we moved to fitting generalised ordered logistic regression models using the user-written **gologit2**, with the “autofit” option allowing the proportional odds assumption to be relaxed for some explanatory variables while being maintained for others^29^. Reducing to parallel one level of a variable separately is consistent with previous approaches^30,31^.

A multivariable generalised ordered logistic regression model was constructed using a backwards stepwise approach to add variables significant at the bivariate stage to the model^32^.

For the second research question, all individuals with an HbA1c indicating diabetes (≥6.5%) were included in the analysis. We chose to focus on this group because individuals with poor glycaemic control, regardless of previous diagnosis, are at increased risk of T2DM complications and are a key population of interest with regard to future public health interventions. Given the large change in sample size for this subset analysis, we started by exploring univariable associations between demographic and socioeconomic factors and healthy diet group, before applying the same steps as for research question one to find the most parsimonious model and investigate whether any variables violated the proportional odds assumption (these steps are shown in appendix 2).

## Results

### Participant Characteristics

The overall sample size of the 2016 SADHS is 10,336 (response rate 81.6%^15^). For the first research question, 3596 participants were excluded from analysis due to not being selected for biomarker sampling, having an invalid HbA1c sample or having an unknown previous diagnosis of diabetes, leaving a sample size of 6709, representing 66% (4159) of female participants and 59% (2581) of male participants^15^.

Table 2 shows the distribution of participants by number of healthy choices and further organisation into healthy diet groups. Most participants demonstrated a mix of healthy and unhealthy dietary choices, albeit with predominantly unhealthy choices. Nearly half of participants had two healthy and three unhealthy choices, with the next largest containing those with one healthy and four unhealthy choices (22.0%). Few participants had an extremely healthy or unhealthy diet with fewer than 1% making five healthy choices and 3.6% five unhealthy choices.

**Table 2.** Distribution of participants by number of healthy dietary choices and organisation into healthy diet groups.

| No. Healthy Choices | Frequency | Percent | Healthy Diet Group |
| --- | --- | --- | --- |
| 0 | 243 | 3.6 | Unhealthy |
| 1 | 1474 | 22.0 | Unhealthy |
| 2 | 3193 | 47.6 | Somewhat Healthy |
| 3 | 1320 | 19.7 | Moderately Healthy |
| 4 | 415 | 6.2 | Healthy |
| 5 | 64 | 0.9 | Healthy |
| Total | 6709 | 100.00 |  |

Figure 1 highlights that across all healthy diet groups in both men and women, low fast food consumption was the dominant healthy choice. Low fruit juice consumption was the dominant unhealthy choice in both men and women across all healthy diet groups. For those in the somewhat healthy diet group (those with two healthy choices), low fast food and low SSB consumption was the most commonly reported combination (54% of all two healthy choice combinations) with this pattern similar in men and women, whilst high fruit juice and high fruit consumption was least commonly reported (4%). For those in the moderately healthy diet group (three healthy choices), low fast food, low SSB and high vegetable consumption was the most commonly reported combination in both men and women (7.5%) whilst high fruit juice, high fruit and high vegetable consumption was least commonly reported in both men and women (0.3%).

**Figure 1:**
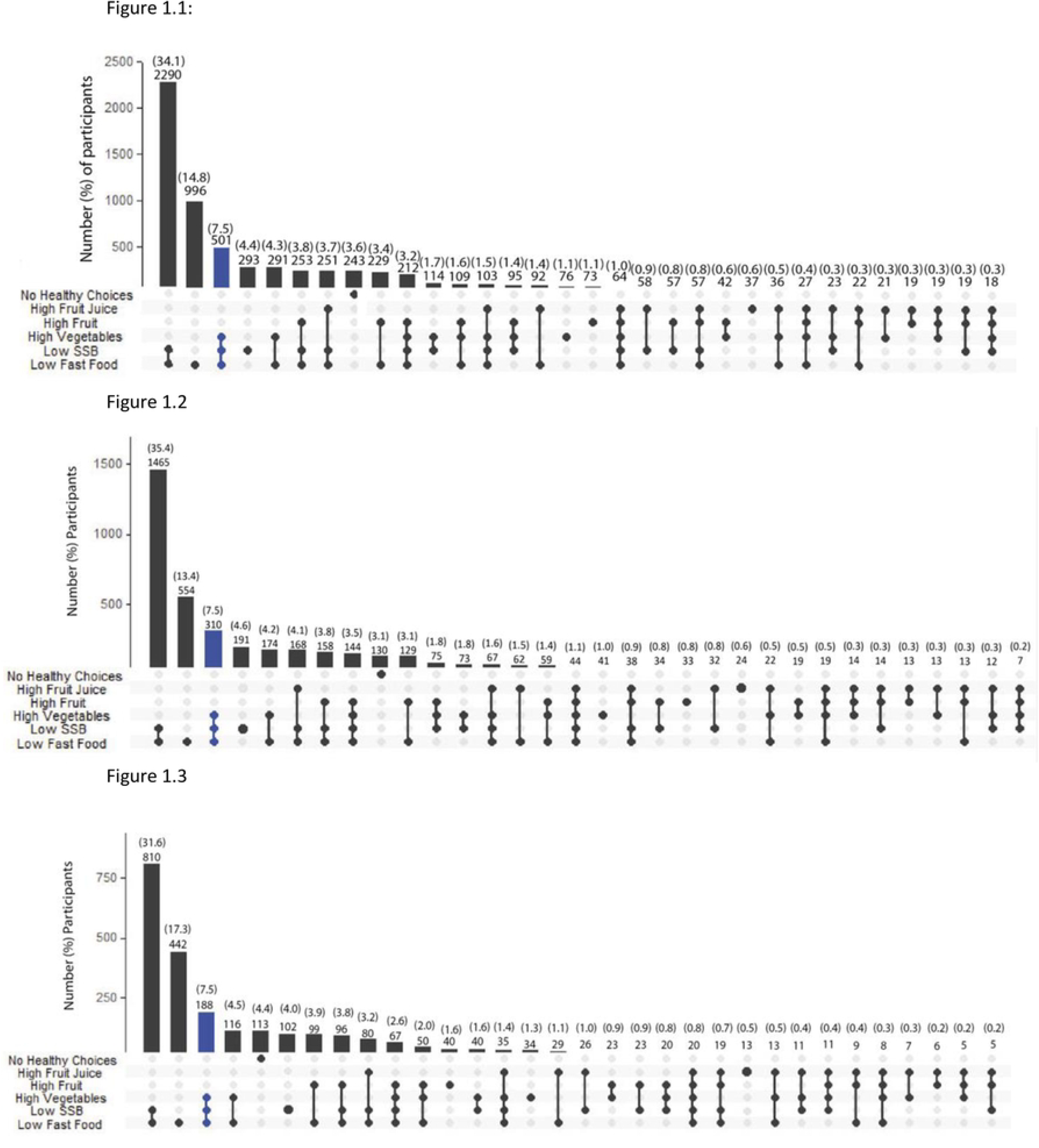
UpSet plots showing patterns of dietary choices among adults aged 15 years and over. <u>Figure 1.1: All adults aged 15 years and over in South Africa (N = 6709)^1,2^</u> ^1^● – For example, a participant with only high vegetable, low SSB and low fast food consumption would be in the moderately healthy diet group. ^2^SSB – sugar-sweetened beverages <u>Figure 1.2: Women aged 15 years and over in South Africa (N = 4148)</u> <u>Figure 1.3: Men aged 15 years and over in South Africa (N = 2560)</u>

Table 3 summarises weighted participant characteristics by healthy diet group, showing that the majority of participants had not previously received a diagnosis of diabetes and that most survey participants had an HbA1c indicating prediabetes. Among those who had received a previous diagnosis of diabetes, 71.9% had poorly-controlled diabetes (HbA1c ≥ 6.5) and a further 25.3% had somewhat-controlled diabetes (5.7% ≤HbA1c ≤ 6.4%). Of those with no previous diagnosis of diabetes, 8.7% had diabetes (HbA1c ≥ 6.5) and 66.3% had prediabetes.

**Table 3.**
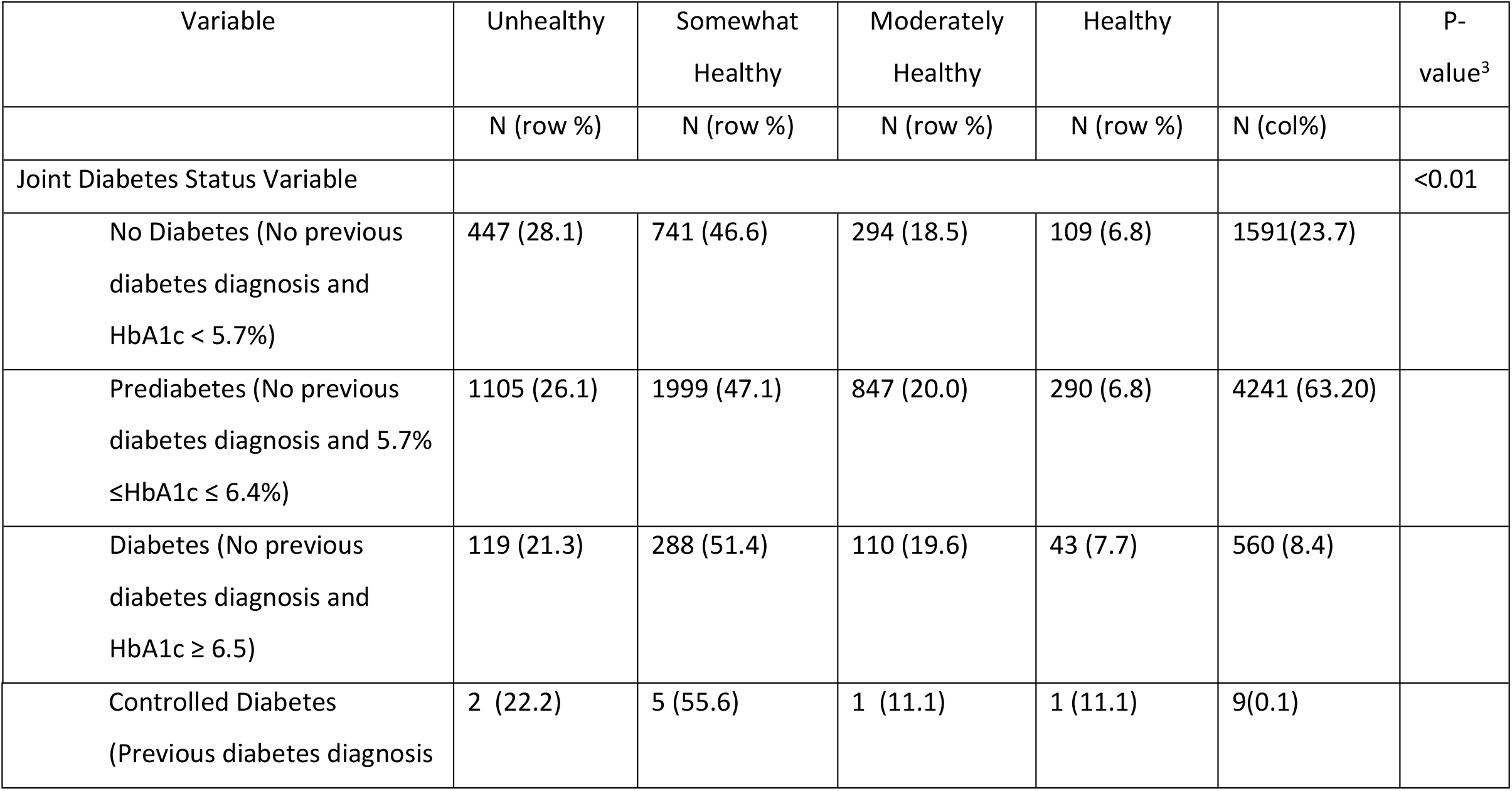
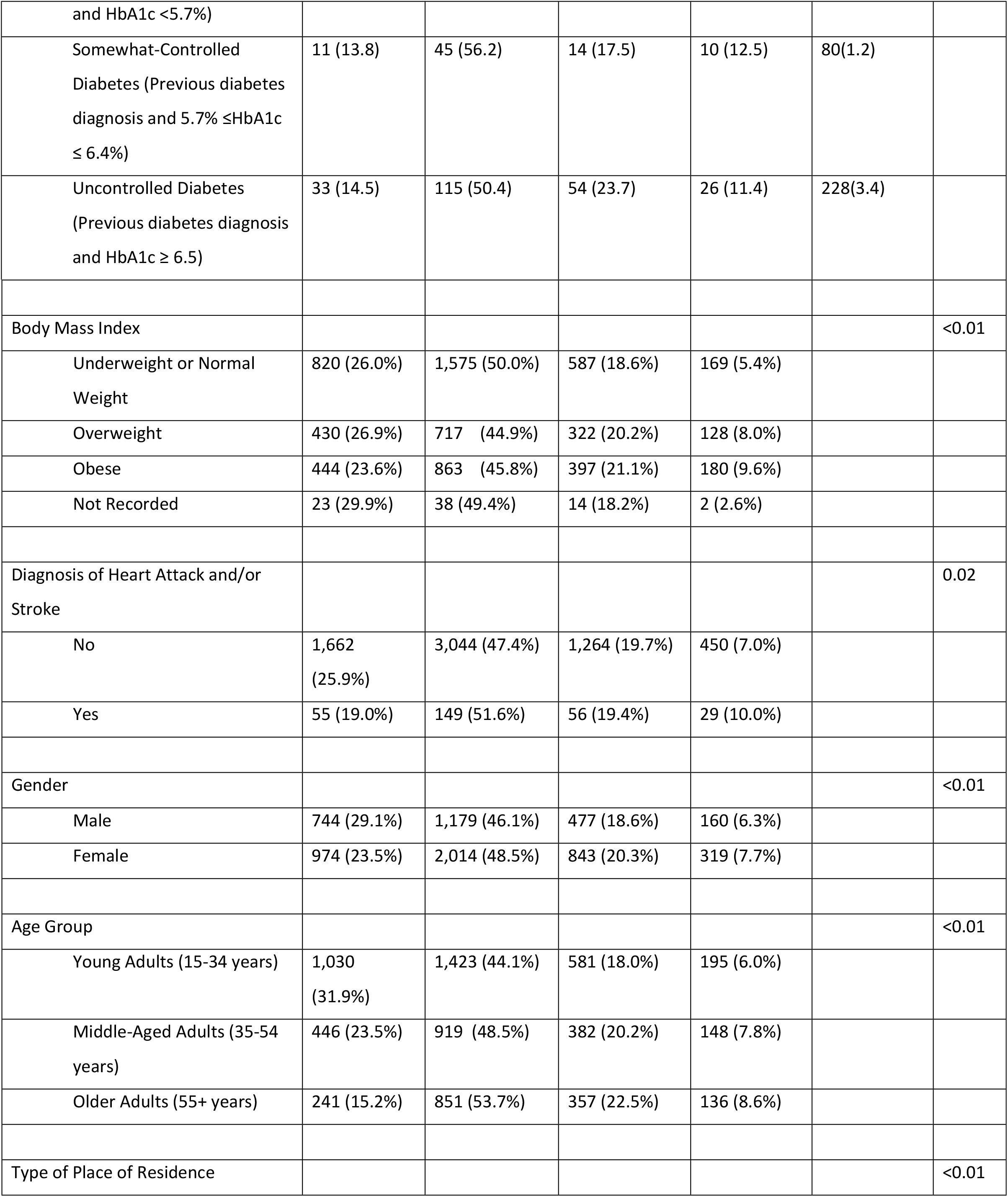
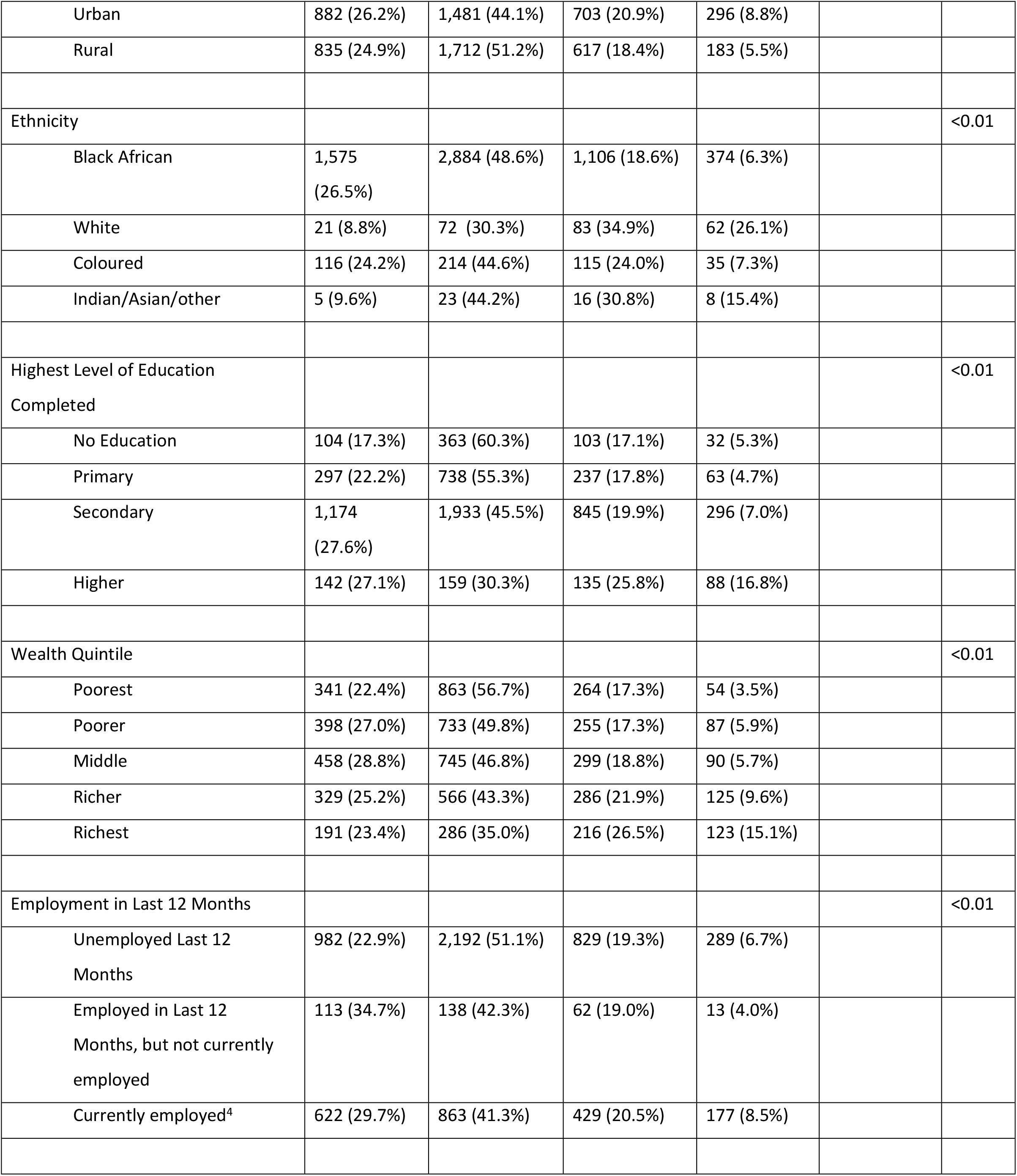
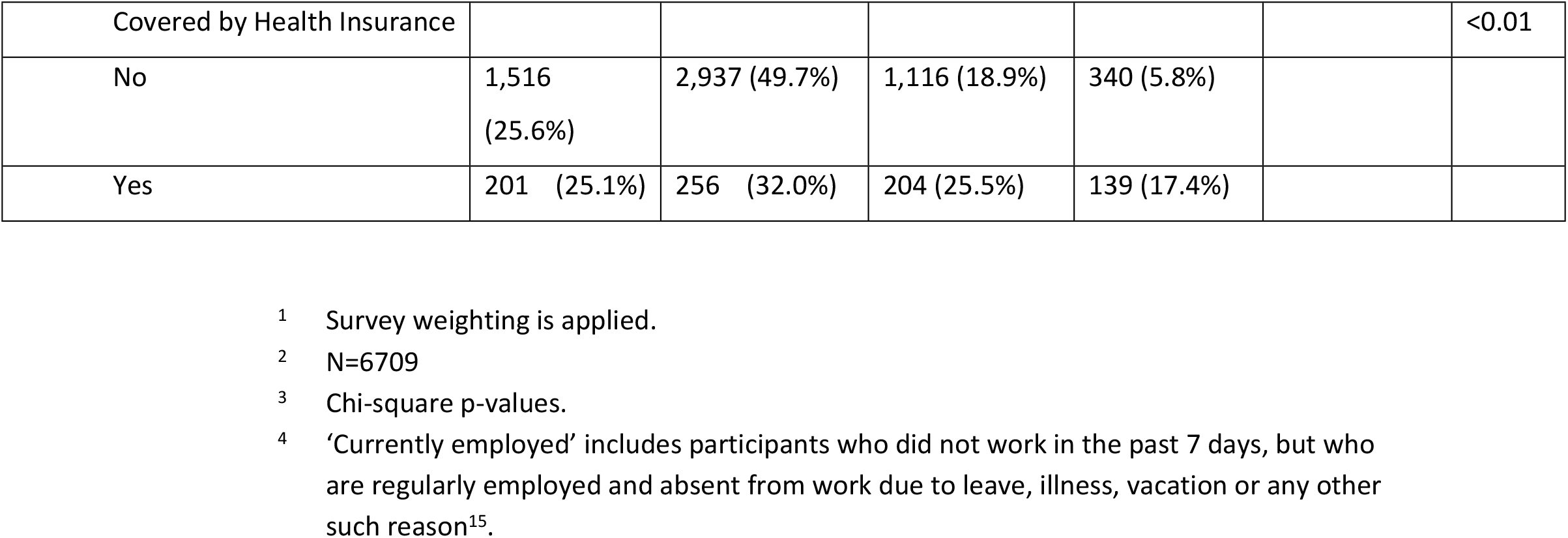
Distribution of Healthy Diet categories by characteristics of survey participants^1,2^.

Appendix 1 shows results of bivariate ordered logistic analyses conducted for research question 1. On bivariate analysis, education showed no pattern of association with diabetes status or healthy diet group and was dropped. Diagnosis of heart attack/stroke was also dropped at this stage as too few individuals reported a diagnosis and it could not be considered for inclusion in multivariate models. As expected, having a previous diagnosis of diabetes was associated with higher odds of being in a healthier diet group relative to individuals without a diagnosis of diabetes (OR 1.83, CI 1.42 – 2.35 for those with uncontrolled diabetes, OR 1.66, CI 1.11 – 2.51 for those with somewhat-controlled diabetes). Among other significant associations, individuals with black African ethnicity had much lower odds of being in a healthier diet group relative to individuals with other ethnicities (OR of being in a healthier diet group 4.43, CI 2.64 – 7.45 for white individuals, OR 1.31, CI 1.01 – 1.69 for coloured individuals and OR 2.53, CI 1.22 – 5.22 for Indian/Asian individuals) and individuals covered by health insurance had higher odds of being in a healthier diet group relative to individuals without health insurance (OR 1.48, CI 1.05 – 1.09).

Likelihood ratio testing of variable contribution at the bivariate stage found that all variables other than type of place of residence and BMI had a p-value<0.10, and were therefore considered in building of the multivariable model. Brant testing of remaining variables at the bivariate stage determined that age, wealth index, employment status in the last 12 months and health insurance coverage violated the parallel odds assumption.

Table 4 shows results of the final multivariable generalised ordered logistic regression model with the joint diabetes status variable, gender, ethnicity and employment in last 12 months reduced to parallel. Individuals with a previous diagnosis of diabetes and uncontrolled diabetes (HbA1c ≥ 6.5) remained significantly more likely to be in a healthier diet group compared to individuals without diabetes (OR 2.14, CI 1.10 – 4.18). No other category of our joint diabetes status variable was statistically significantly different in odds of a healthier diet compared to the base group with no diabetes (no previous diagnosis and an HbA1c<5.7%), although the estimate for the somewhat controlled is in the same direction while numbers contributing to the controlled diabetes group estimate are very small (N=18). Middle-aged and older adults had significantly greater odds of being in the somewhat healthy, moderately healthy or healthy diet groups than the unhealthy diet group relative to younger adults (OR 1.54, CI 1.28-1.86 for middle aged adults; OR 2.06, CI 1.23-3.43 for older adults). This association is attenuated and becomes statistically insignificant when comparing individuals in the unhealthy and somewhat healthy diet groups to those in the moderately healthy and healthy diet groups by age (OR 1.04, CI 0.86 – 1.27 for middle aged adults, OR 1.22, CI 0.77 – 1.95 for older adults).

**Table 4.**
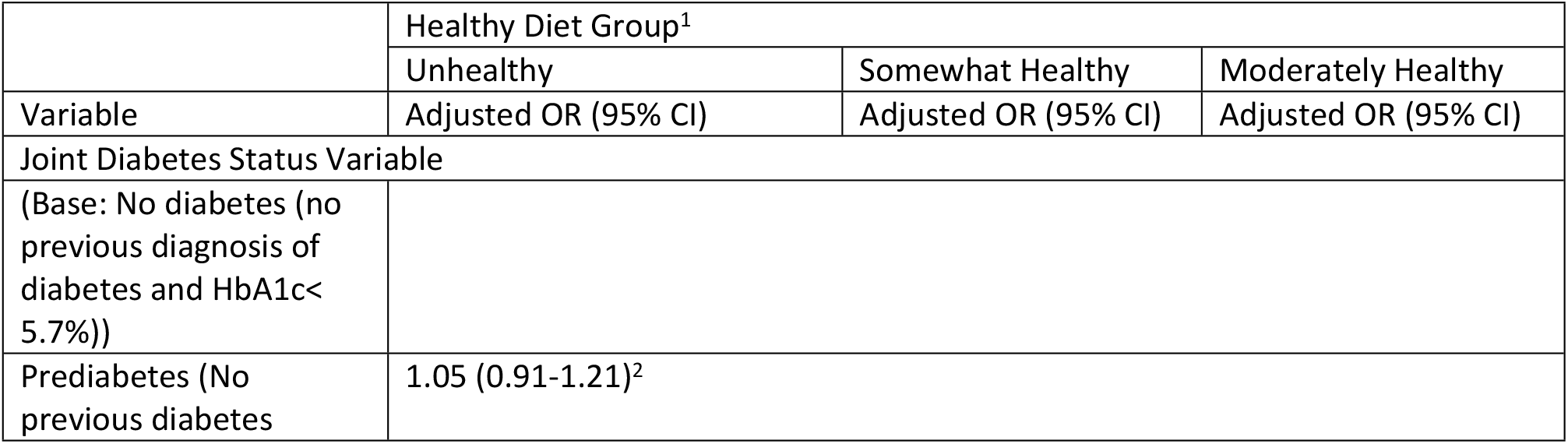
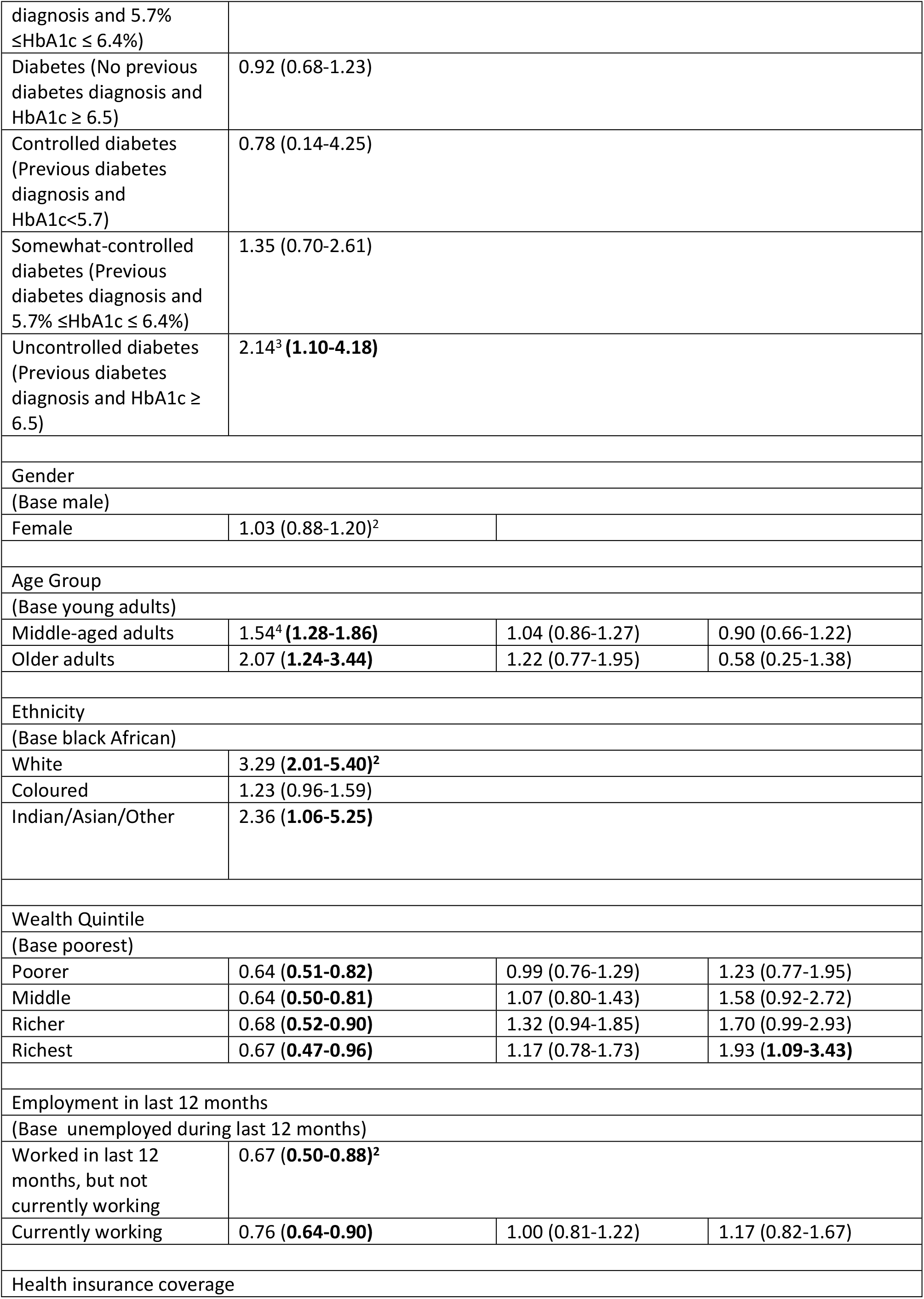
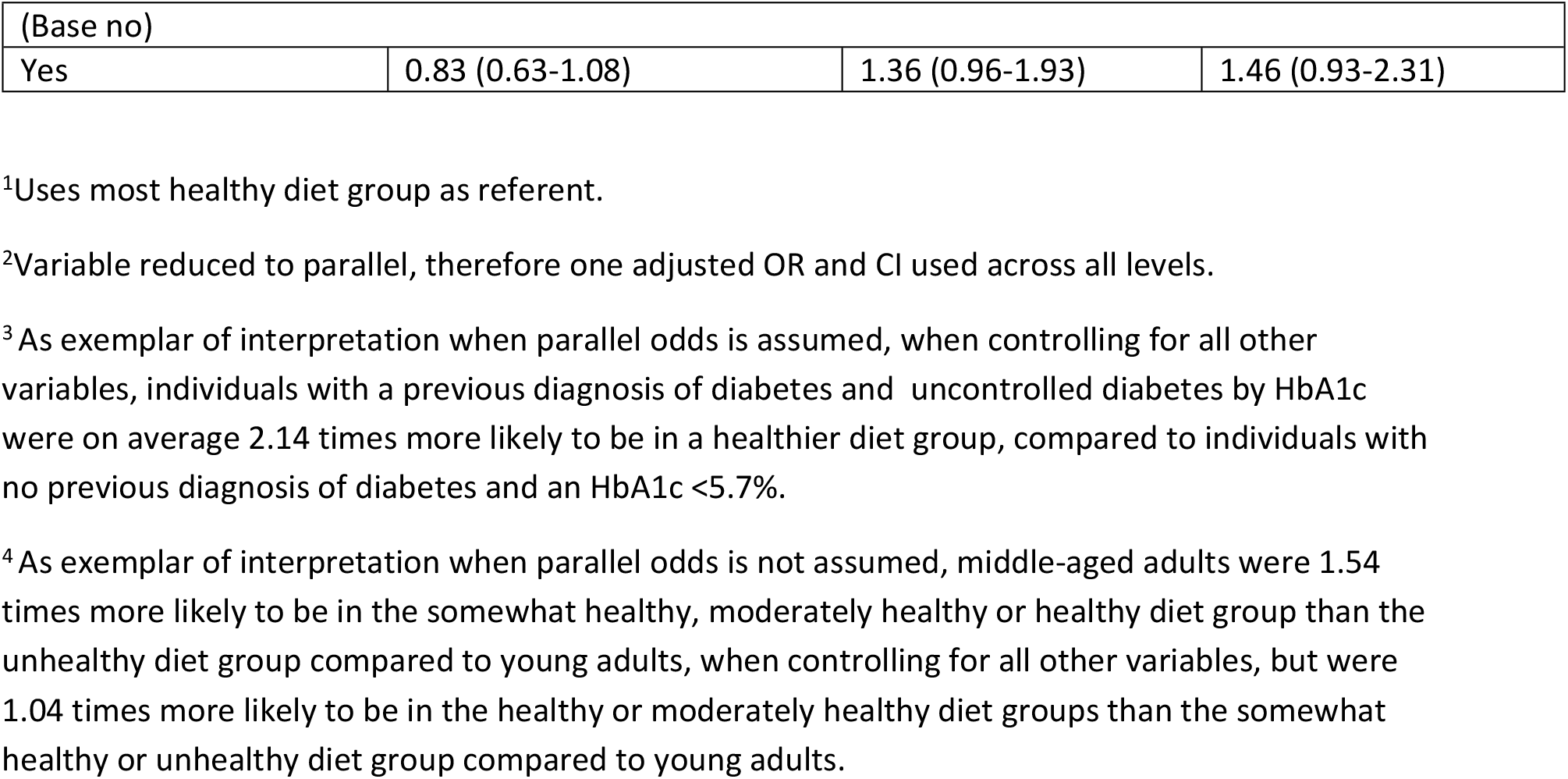
Final multivariable generalised ordered logistic regression model for the odds of healthy diet.

Individuals with black African ethnicity remained significantly less likely to have healthy diet than all other ethnicities. There was a u-shaped relationship between wealth quintile and diet, with all wealth quintiles more likely to be in the unhealthy diet group than the healthy, moderately healthy or somewhat healthy diet groups relative to the poorest quintile, but higher wealth quintiles having an increasingly higher likelihood of being in the healthy diet group than the moderately healthy, somewhat healthy or unhealthy diet group compared to the poorest quintile, with this being significant for the richest wealth quintile (OR 1.95, CI 1.10-3.46). Individuals who were employed in the last 12 months but were currently unemployed were significantly less likely to have a healthy diet relative to those unemployed throughout the last 12 months (OR 0.66, CI 0.50-0.88), whilst those currently employed were significantly more likely to be in the unhealthy diet group (OR 0.76, CI 0.64-0.90).

### People Living With Diabetes

The distribution of individuals with uncontrolled diabetes defined by HbA1c, irrespective of previous diagnosis, was broadly similar to in the general population, with the exceptions that individuals with three healthy and two unhealthy choices were the second largest group rather than the third largest among PLWD. Overall, the distribution of PLWD by sociodemographic status was similar to that in the general population. This can be further seen in appendix 2.

Likelihood ratio testing found that employment status, education status, ethnicity and type of place of residence had a p-value>0.10 and these variables were dropped. Further likelihood ratio testing was conducted on smaller models in a backwards stepwise manner as for research question one. Age, gender, wealth quintile and health insurance coverage were retained for further analyses after this testing. Brant testing of the final ordered logistic regression model showed that none of the included variables violated the parallel odds assumption, meaning the ordered logistic regression model was retained, as seen in appendix 3.

Table 5 shows results from the final ordered logistic regression model of healthy diet among PLWD. When controlling for other variables, females living with diabetes were significantly more likely than males to be in a healthier diet group (OR 1.40, CI 1.03-1.90). Middle-aged adults were no more likely than younger adults to be in a healthier diet group, however older adults were borderline significantly more likely to be in a healthier diet group than younger adults, (OR 1.55, CI 0.99-2.43). Those in the richer (OR 1.63, CI 1.06-2.51) and richest (OR 1.85, CI 1.14-3.02) quintiles were significantly more likely to be in a healthier diet group relative to the poorest group. Health insurance coverage, holding wealth quintile and all other factors constant, showed an independent statistically significant association with healthy diet group - those with coverage were almost twice as likely as those without coverage to have a healthier diet (OR 1.96, CI 1.27-3.01).

**Table 5.**
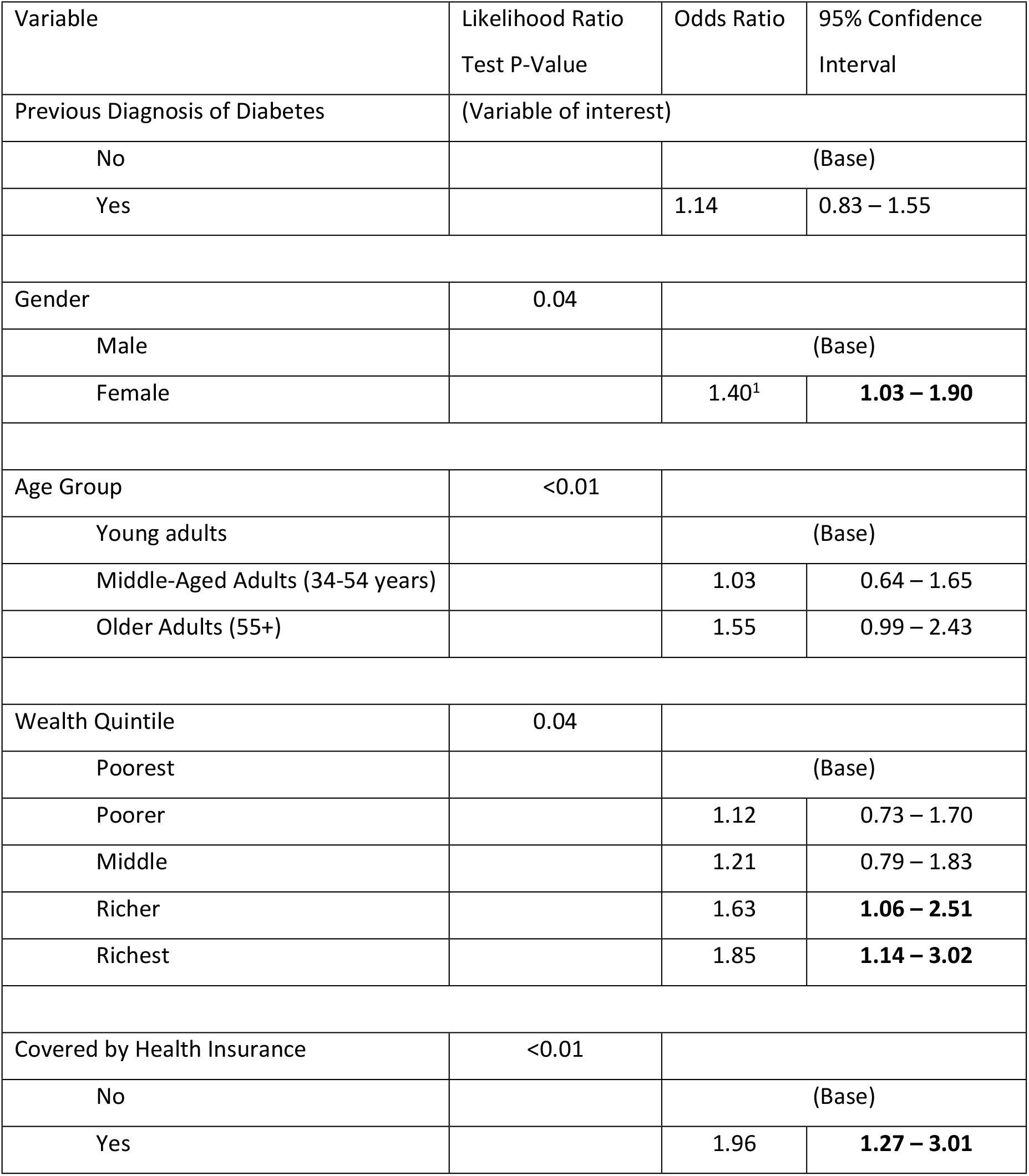
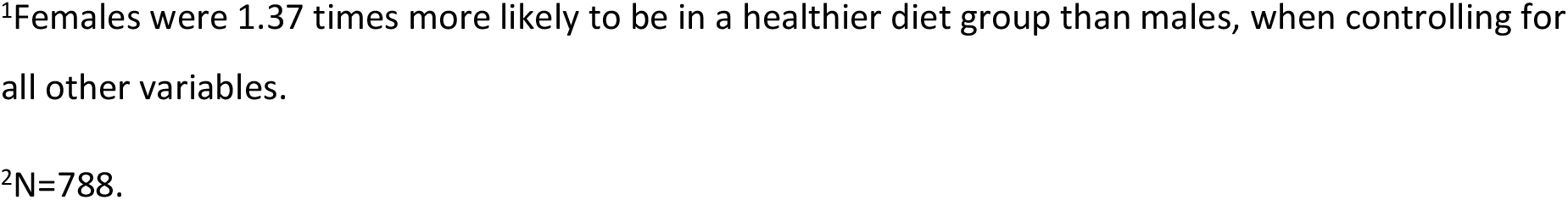
Final Multivariable ordered logistic regression model of healthy diet among people living with diabetes^1,2^.

| Variable | Likelihood Ratio<br>Test P-Value | Odds Ratio | 95% Confidence<br>Interval |
| --- | --- | --- | --- |
| Previous Diagnosis of Diabetes | (Variable of interest) |  |  |
| No |  | (Base) |  |
| Yes |  | 1.14 | 0.83 – 1.55 |
| Gender | 0.04 |  |  |
| Male |  | (Base) |  |
| Female |  | 1.40<sup>1</sup> | **1.03 – 1.90** |
| Age Group | <0.01 |  |  |
| Young adults |  | (Base) |  |
| Middle-Aged Adults (34-54 years) |  | 1.03 | 0.64 – 1.65 |
| Older Adults (55+) |  | 1.55 | 0.99 – 2.43 |
| Wealth Quintile | 0.04 |  |  |
| Poorest |  | (Base) |  |
| Poorer |  | 1.12 | 0.73 – 1.70 |
| Middle |  | 1.21 | 0.79 – 1.83 |
| Richer |  | 1.63 | **1.06 – 2.51** |
| Richest |  | 1.85 | **1.14 – 3.02** |
| Covered by Health Insurance | <0.01 |  |  |
| No |  | (Base) |  |
| Yes |  | 1.96 | **1.27 – 3.01** |

<sup>1</sup>Females were 1.37 times more likely to be in a healthier diet group than males, when controlling for all other variables.
<sup>2</sup>N=788.

## Discussion

Having both a previous diagnosis of diabetes and an HbA1c at the time of survey indicating uncontrolled diabetes was associated with a healthier diet, whilst having an HbA1c indicating diabetes, but no previous diagnosis of diabetes was not associated with healthy diet group after controlling for other variables. Given the role of diagnosis as an opportunity for discussing, facilitating and motivating lifestyle change^23^, this difference illustrates that the large number of individuals with undiagnosed diabetes are at increased risk of continuing to make unhealthy dietary choices. Although the majority of individuals with a diagnosis of diabetes still had poor glycaemic control, it may be the case that diet change alone was insufficient to induce glycaemic control in this group, whilst care cascade data reported that almost half of individuals receiving treatment still have uncontrolled diabetes (HbA1c≥6.5%)^3^. Existing literature shows that black ethnicity and low socio-economic status (SES) are predictors of undiagnosed diabetes^26^ and poor healthcare access^33^, whilst our results also show they are associated with unhealthy diet among both PLWD and the general population. This means PLWD in sociodemographic groups more likely to have unhealthy diet prior to diagnosis of diabetes are also less likely to have access to healthcare professionals who can help to motivate lifestyle change, in addition to being economically less able to introduce them, placing this group at increased risk of T2DM complications. Encouragingly, early NCD detection in primary care settings is included as a strategic action area in the new 2020-2025 NCD plan created by the South African Department of Health (SADOH)^34^. Planned strategies include the development and review of a diabetes cascade system to mirror the structure used for HIV management, training community health workers to conduct diabetes screening and advocating for T2DM screening to be covered by national health insurance. Integrating T2DM screening with improved provision of lifestyle counselling and education in primary care widens access to appropriate diabetes diagnosis and management and may bring about healthier dietary choices among PLWD.

The pattern of concurrent low fruit, fruit juice and vegetable consumption seen among both the general population and PLWD is a common issue observed in South African population studies^11,25,35^. Although these studies are limited by their cross-sectional nature, longitudinal data from a repeat panel study by Ronquest-Ross et al^36^ found low fruit and vegetable consumption at baseline and showed a 7.9% reduction in vegetable consumption from 42.0 to 38.7 kg/capita/year, albeit a 6.4% increase in fruit consumption 28.1 to 29.9 kg/capita/year between 1999 and 2012. Although low fruit and vegetable intake is a global problem^36^, it is of particular concern in South Africa and other LMICs due to the rapid and unplanned nature of urban population growth^12^, general increases in the cost of living^37^ and unemployment^38^, and rapidly rising levels of NCDs against which fruit and vegetables are protective. Although this study found low rates of SSB consumption, their intake among the general population in South Africa increased by 68.9% from 55 L/capita/year to 92.9 L/capita/year between 1994 and 2012^36^.

Analysis in both the general population and PLWD found an association between older age and healthier diet group. Older age has previously been associated with higher fruit and vegetable consumption^25^ and qualitative research has suggested that older populations may have a greater preference for vegetables relative to younger individuals^39^. In contrast with these results, research involving PLWD recruited at hospital clinics found no association between age and healthier diet^40^. Given that NCDs seen primarily in older individuals are already increasing in prevalence, evidence that dietary quality is decreasing in younger age groups, combined with the high rate of prediabetes seen in this study, is concerning as it confirms predictions^1,2^ that the burden of T2DM will continue to grow without urgent action.

This study and more localised research^40^ found that women with T2DM had a healthier diet than men. These findings also fit into wider patterns of demographic transition in South Africa which show that men have an increasingly more equal risk of obesity^41^, as well as a greater chance of being physically inactive compared to women^42^, and point towards the future burden of NCDs becoming higher among men, as in high-income countries^2^, with females having lower albeit still burdensome rates of complications.

Despite historic links between ethnicity and wealth in South Africa, we found that lower wealth and black African ethnicity were both independently associated with a less healthy diet. This suggests that the socioeconomic effects of apartheid extend further than wealth alone, but also impact individuals’ food environment. Previous literature has shown that those with lower wealth have poorer dietary diversity^40^, as well as reduced fruit and vegetable consumption^25^. Qualitative and quantitative studies have proposed that this is due to the high cost of fresh fruit and vegetables^11,39^, with affordability the most important factor for South Africans when making dietary choices^34^. Our results additionally noted an increased likelihood of having unhealthy diet for those in higher wealth quintiles relative to the poorest, which could be explained by the inclusion of fast-food consumption in our models, which is higher among those with white ethnicity and higher SES^10^. This would indicate that unlike in high-income countries, where those in more deprived areas have greater exposure to fast-food and takeaway outlets^43^, fast-food is currently more popular among South Africans with higher SES.

These findings, as well as existing evidence, suggest that future public health interventions and policies should focus on making fruit and vegetables more accessible, particularly to younger, majority black populations with low SES. The SADOH NCD plan includes the intention to make healthy foods more available and affordable, but identifies few specific target populations or policies. Participation in a previous intervention giving 25% cashback on healthy dietary purchases was associated with an increase in fruit and vegetable consumption and a decrease in sugary and fast-foods^43^, but was only available to members of a private health insurance company and therefore targeted a demographic already more likely to have the means to purchase healthier foods. Targeted government-funded interventions have so far only focused on providing nutritious meals and health education for school age children^44^, although these interventions do use schools as a conduit for educating parents and families. Whilst this is valuable given the rising rates of childhood obesity reported recently^7^, qualitative evidence shows that people in low-income areas are often aware of the implications of unhealthy diet^39,45,46^ and more work is therefore needed to create a more enabling food environment in such areas. Research has also documented overly simplistic, and even shaming, advice from healthcare professionals in regard to improving diet^46^ and there is a need for additional recognition of environmental barriers when motivating lifestyle change in healthcare settings. A tax on SSBs was recently introduced in South Africa alongside a levy on fruit juice, with SSBs subsequently increasing in price^47^. Whilst this could serve as a model for similar policies on foods, no relative price decrease was detected for healthier beverages and so more far-reaching policies may be required to reduce the cost and increase availability of healthy foods and avoid exacerbating food insecurity among low SES households.

Whilst South Africa’s population sub-groups and history make racial and socioeconomic inequalities particularly entrenched, these findings remain increasingly generalisable to other LMICs as the global epidemiological and nutritional transitions continue. Obesity rates in all Southern Africa Development Community Countries increased between 1990 and 2019^7^, whilst rates of urbanisation in LMICs are also rising more rapidly than in high-income countries^13^. This indicates that many of the challenges faced by South Africa will also be faced by other LMICs in future years and whilst morbidity and mortality attributed to T2DM remains lower in other areas of sub-Saharan Africa^1^, rates are beginning to increase. Proactive strategies for prevention and treatment of T2DM will therefore be needed in many LMICs to address the future burden of disease, including those addressing dietary risk factors.

### Strengths and Limitations

The large, nationally representative sample size of the SADHS is a strength of this study, as is the number of available variables, which allows for consideration of multiple dietary groups and potential confounding factors. Using Hba1c as the primary measure of diabetes status rather than self-reported diagnosis is biologically objective and enables the inclusion of individuals with undiagnosed diabetes, although it is an indirect measure of blood glucose and utilises only one measurement, making it less reliable than a fasting glucose test^48^. The use of generalised ordered and ordered logistic regression models the hierarchical nature of healthy diet groups. To the authors’ knowledge, this is the first study to use such modelling to investigate the associations between diabetes status and diet.

The cross-sectional nature of the SADHS is a limitation and means it was not possible to assess changes over time. Four out of five dietary variables only considered consumption during the previous day, limiting the representativeness of the survey as a consistent measure of individual dietary choices. The low number of individuals in the healthy diet group limited statistical power when comparing with this group, whilst the use of a nationally representative sample meant that sample sizes for minority groups, such as the Indian/Asian and elderly populations were small. Given the low number of individuals reporting fruit juice consumption, including this with fruit and vegetable consumption to create a binary variable of 5+ vs <5 portions of fruit and vegetables per day (reflecting national recommendations) may have improved statistical power. Other variables associated with healthier dietary outcomes in the literature, such as increased grocery expenditure^25^ and increased time spent cooking^43^, were not collected in the 2016 SADHS and could not be considered in these analyses. Whilst the prevalences of other types of diabetes mellitus in South Africa are very small relative to T2DM^1,49^, the SADHS does not differentiate between types of diabetes.

Our findings, in combination with existing evidence, should inform the development of actionable public health policies in South Africa, with a particular focus on improving fruit and vegetable consumption in younger, black and low-SES populations without access to health insurance clearly required. Key healthcare stakeholders including the South African government, non-government organisations and health insurance providers should integrate T2DM screening with individualised lifestyle management, as well as population-level interventions, to combat the rising burden of the disease.

## Data Availability

Data cannot be shared publicly as per DHS program terms and conditions of use. SADHS 2016 datasets are available from the DHS program website (https://www.dhsprogram.com/data/Access-Instructions.cfm) upon request.

## List of Abbreviations

PLWD: person/people living with diabetes.
LMIC: low or middle-income country
NCD: non-communicable disease
T2DM: Type 2 diabetes mellitus
SADHS: South African Demographic and Health Survey
HbA1c: Glycosylated haemoglobin
EA: Enumeration area
PSU: Primary sampling unit
SSB: Sugar-sweetened beverage
UPF: Ultra-processed food
BMI: Body mass index
SADOH: South African Department of Health
SES: Socioeconomic status

## Declarations

### Human Ethics and Consent to Participate

Ethical approval was received from the Faculty of Medicine Ethics Committee, University of Southampton, under approval number 66804.

Participants gave informed verbal consent to give survey data and informed written consent to give biomarker data to the SADHS. All data used in this paper is anonymised. For participants aged 15-17, consent was required from both the participant and their legal parent/guardian.

All research was conducted in accordance with the Declaration of Helsinki.

### Consent for Publication

Not applicable

### Author’s Contribution

MB was responsible for study design and data analysis under guidance from NM. Survey and biomarker data was collected by the Demographic and Health Survey program, who gave approval to the authors for use in this study.

## Acknowledgements

The authors would also like to thank Dr Beth Stuart for some additional guidance of statistical model choice and Dr Zhixin Feng for guidance on variable weighting.

